# Altered neurodevelopmental trajectories of brain structure in Tourette syndrome and Chronic Tic Disorders

**DOI:** 10.64898/2026.05.16.26353368

**Authors:** Yin Jin, Yuxin Guo, Jonathan M. Koller, Sarah C. Grossen, Anne Uhlmann, Natalie J. Forde, Jade-Jocelyne Zouki, Renzo Torrecuso, Karsten Mueller, Juan F. Martin-Rodriguez, Pablo Franco-Rosado, Michel Grothe, Catharina Cramer, Anne Kleine Büning, Heike Eichele, Stefano Palmucci, Adriana Prato, Federica Saia, Silvia Tommasin, Giulia Conte, Katharina A. Schindlbeck, Christos Ganos, Shukti Zimmermann, Tanja Veselinovic, Yulia Worbe, Andreas Hartmann, Apostolia Topaloudi, Mary Kaka, Guangxin Chen, Qingyi Zhong, Yuqi Zhang, Natalia Szejko, Piotr Janik, Nanette M. Debes, Zeynep Tumer, Tomasz Wolańczyk, Gary A. Heiman, Hreinn Stefansson, Helga Ask, Ole Andreassen, Anders D. Børglum, Joseph D. Buxbaum, Elizabeth C. Corfield, Mark Daly, Dorothy E. Grice, David Mataix-Cols, Aarno Palotie, Christian Rück, Matthew W. Halvorsen, Lea K. Davis, James J. Crowley, Manuel Mattheisen, Dongmei Yu, Carol A. Mathews, Jeremiah M. Scharf, The members of the Tourette International TS-EUROTRAIN/TSGeneSEE, TAA Neuroimaging Consortium, Collaborative Genetics consortium, TAAICG, EMTICS, ENIGMA-TS Working Group, Dan J. Stein, Irene Neuner, Richard Musil, Francesco Cardona, Danielle Cath, Dick J. Veltman, Tim J. Silk, Alexander Munchau, Julius Verrel, Valerie C. Brandt, Tamara Hershey, Deanna J. Greene, Bradley L. Schlaggar, Jan Buitelaar, Barbara Franke, Sophia Thomopoulos, Renata Rizzo, Andrea Dietrich, Pieter J. Hoekstra, Pablo Mir, Veit Roessner, Odile A. van den Heuvel, Kirsten Mueller-Vahl, Harald Möller, Neda Jahanshad, Paul M. Thompson, Kevin J. Black, Peristera Paschou

## Abstract

Tourette syndrome (TS) is a neurodevelopmental disorder characterized by symptoms that emerge in childhood and often improve or even disappear in adulthood, providing a model for understanding how altered brain development shapes neural structure and function. We investigate brain structural alterations in TS and Chronic Tic Disorders (TS/CTD) across development, presenting the largest structural neuroimaging analysis for TS/CTD to date (1,803 individuals from the ENIGMA-TS Working Group), and integrating with large-scale genomewide association studies. Nonlinear age effects were observed in cortical thickness across development and in thalamic volume in children, indicating altered trajectories of brain maturation. Pediatric and adult TS/CTD showed distinct structural patterns, with widespread alterations in childhood and more focal changes in adulthood. Children also showed the most prominent effects highlighting the involvement of orbitofrontal cortex and putamen, alongside additional regions such as frontal and paralimbic areas. Genetic pleiotropy analyses identified overlap between TS/CTD-associated genetic effects on brain structure and neuroanatomical differences. Cross-disorder comparisons revealed correlations with ADHD and OCD and age-related patterns. These findings demonstrate altered neurodevelopmental trajectories in TS/CTD and implicate systems underlying inhibitory control and urge regulation.

---

Tourette syndrome (TS) is a neurodevelopmental disorder with a lifetime prevalence of approximately 1% and is characterized by motor and vocal tics that emerge in early childhood, peak in late childhood or early adolescence, and often improve or even disappear in adulthood ^1,2^. Chronic Tic Disorders (CTD) follow a similar pattern, and only approximately 20% of children with CTD retain their diagnoses in adulthood ^3,4^. This characteristic trajectory provides a model for investigating how altered brain development across the lifespan shapes neural structure, function, and behavior. TS and CTD (TS/CTD) are also often comorbid with other neurodevelopmental disorders, and most frequently with attention-deficit/hyperactivity disorder (ADHD; up to 54%), and obsessive-compulsive disorder (OCD; up to 50%), suggesting shared underlying mechanisms ^5,6^. TS/CTD are highly heritable ^7^, and genome-wide association studies (GWAS) have implicated pathways related to ligand-gated ion channel signaling, immune processes, cell adhesion, and synaptic signaling ^8–12^, primarily converging on cortico–striato– thalamo–cortical (CSTC) circuits in the most recent TS/CTD GWAS ^13^. However, despite these advances, how genetic risk and neurodevelopmental processes interact to shape brain structure from childhood to adulthood in TS/CTD remains poorly understood.

Structural magnetic resonance imaging (sMRI) studies of TS/CTD have also provided important insights that complement genetic approaches. However, they have been constrained by small sample sizes and substantial heterogeneity, resulting in inconsistent findings that have hindered definitive biological interpretation. In concordance with recent genetic studies, CSTC circuits have been implicated ^13^, with case–control differences reported predominantly in the basal ganglia ^14–17^, thalamus ^18,19^, and prefrontal cortex (PFC) ^20–24^. However, previous findings include both increased and decreased volumes of the putamen and thalamus ^14–19^ as well as inconsistent results in prefrontal regions ^20–24^. Such discrepancies likely reflect limited sample sizes, variation in imaging acquisition and analytical approaches, and differences in the age composition of study samples. This issue is particularly relevant for TS/CTS, with symptoms and potential compensatory processes evolving from childhood to adulthood. Previous studies suggest that brain alterations may differ across age groups, including abnormal PFC volumes reported in children but not adults ^24^ and opposite effects on insular cortex morphology across developmental stages ^25,26^. Functional MRI (fMRI) studies further support involvement of CSTC networks by demonstrating altered connectivity among these regions ^27–29^ and indicate that patterns may vary with age ^30^. Together, these findings highlight the need for large, harmonized neuroimaging studies capable of providing robust characterization of brain structure in TS/CTD in different age groups.

Given the high percentage of other neurodevelopmental disorders as comorbid in TS/CTD, large-scale neuroimaging studies are also essential for disentangling disorder-specific and shared neural alterations. Previous studies have shown partial neuroimaging overlap of TS/CTD with other neurodevelopmental disorders, including ADHD, OCD, and ASD, particularly within basal ganglia and frontal regions ^17,31–41^. Although all these conditions share involvement of cortico– subcortical networks, previous studies suggest that each disorder also shows distinct neuroanatomical profiles ^17,31–41^. Cross-disorder analyses from the ENIGMA (Enhancing Neuroimaging Genetics through Meta-analysis) consortium, comparing ADHD, OCD, and ASD ^35^, further demonstrated disorder-specific and age-dependent patterns of structural brain differences. These findings further underscore the need for harmonized large-scale studies for TS/CTD, capable of disentangling disorder-specific and transdiagnostic effects.

Here, we leverage the ENIGMA-TS (Enhancing Neuroimaging Genetics through Meta-analysis for TS) Working Group ^42^ to characterize structural brain alterations in TS/CTD across development and to examine how these differences vary with age, providing insight into altered neurodevelopmental trajectories. In doing so, we address heterogeneity in previous neuroimaging findings, analyzing the largest coordinated structural neuroimaging dataset assembled for TS/CTD to date. We recruited regional volumetric measures derived from T1-weighted structural MRI scans from 1,803 participants across 13 sites from 8 countries using standardized ENIGMA processing and quality control pipelines ^43^. We conducted complementary mega- and meta-analyses to identify cortical and subcortical structural differences associated with TS/CTD and tested age-by-diagnosis and sex-by-diagnosis interactions to characterize developmental effects. To further explore biological mechanisms, we performed genetic pleiotropy analyses linking TS/CTD polygenic risk to brain structure and compared the resulting neuroanatomical patterns with those observed in ADHD and OCD, the most common comorbid conditions in TS/CTD. By integrating large-scale neuroimaging, genetic, and cross-disorder perspectives, this study provides the most comprehensive characterization to date of structural brain alterations in TS/CTD from childhood to adulthood and offers new insight into how genetic risk and neurodevelopment may converge to shape the clinical expression of tic disorders.

## Methods

### Datasets

Leveraging the ENIGMA-TS Working Group ^42^ and standardized ENIGMA pipelines for harmonized multisite analysis, we analyzed a dataset comprising N = 1,803 individuals who underwent T1-weighted structural MRI (T1w sMRI) across 13 data providing sites in 8 countries using locally established acquisition protocols (see Supplementary Table S1 and Figure S1 for site and country information). Diagnoses of TS and CTD, as well as comorbid ADHD and OCD, were established according to DSM-5 diagnostic criteria.

After a rigorous quality control (QC) pipeline (see Supplementary Methods and Supplementary Figure S2), 1,648 participants aged 2–68 years were retained for analysis, including 745 cases (725 with TS and 20 CTD, here collectively referred to as TS/CTD) and 903 controls. Participants were grouped into 21 cohorts according to the contributing site, scanner characteristics, and age composition of the dataset. To account for age-related effects on diagnostic differences that were observed in our interaction analyses, analyses were also conducted separately for pediatric (<18 years) and adult (≥18 years) groups. More details are provided in Supplementary Table S2. The study adhered to all local institutional review board guidelines for the ethical use of coded data in research.

### Image Acquisition and Processing

T1w brain sMRI scans were collected independently across sites and processed following the standardized ENIGMA pipeline protocols (see also supplementary material) ^43^, as broadly implemented previously (https://enigma.ini.usc.edu/protocols/imaging-protocols/). Details of image acquisition parameters for each cohort are available in Supplementary Table S1. All T1w brain sMRI scans were analyzed using FreeSurfer software (version 7.0 and above) ^44^, enabling consistent analysis and quality control (see supplementary material) across all participating sites. Measurements of surface area and cortical thickness were extracted for 68 cortical regions, with 34 regions in the left hemisphere and 34 in the right hemisphere, according to the Desikan-Killiany atlas ^45^. Additionally, average thickness and total surface area were calculated per hemisphere. 16 subcortical volumes, including 14 bilateral subcortical grey matter volumes (nucleus accumbens, amygdala, caudate, hippocampus, pallidum, putamen, and thalamus) and the two lateral ventricles, were extracted using all segmented areas using the ASEG atlas ^46^. QC procedures involved both visual inspections and statistical evaluations, including the use of Cook’s distance to identify and manage outliers ^47^.

### Comparison of Brain Structure Between TS/CTD and controls: Mega- and Meta-Analyses

We performed a mega-analysis to investigate structural brain differences between individuals with TS and controls in the 68 cortical and 16 subcortical regions of interest (ROI) in the entire sample as well as separately in the pediatric and adult groups. Following standard pipelines from prior ENIGMA studies ^34,39,48^, linear mixed-effects modeling was chosen for our primary analysis. This is a widely established method for multisite neuroimaging data analysis, effectively adjusting for mean site differences by modeling site-specific means as random intercepts ^49^. The model was implemented in R (4.4) with the *lme4* package ^50^, and it included fixed effects of age, squared age (age^2^), sex, intracranial volume (ICV) (for surface area and subcortical volumes), and interactions between age and sex, as well as random intercepts to account for cohort variation (see equation 4.1). False discovery rate (FDR-adjusted p) of < 0.05 as the criterion for statistical significance was applied. Effect sizes were calculated using Cohen’s *d* ^*51*^, derived from t-values from the mixed-effects model. To evaluate the impact of age and sex on TS diagnosis, we also included age-by-diagnosis and sex-by-diagnosis interaction terms in our models and applied FDR correction in subsequent analyses. For consistency with the prior ENIGMA studies ^48,52,53^, a meta-analysis was also conducted and assessed its agreement with the primary mega-analysis using Pearson correlations. Further methodological details are available in the supplementary materials.

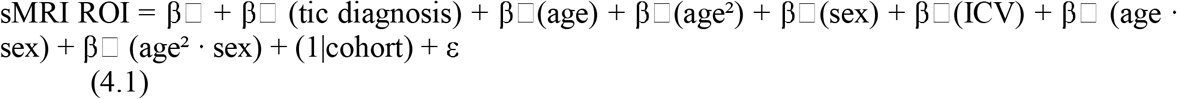

### Sensitivity Analysis Controlling for Comorbid ADHD, OCD, and Brain-Active Medication Use and Batch-effect

A series of sensitivity analyses were conducted to examine whether brain structural differences associated with TS are robust to the confounding effects of comorbid ADHD, OCD, and brain-active medication use, given their high prevalence in TS/CTD (for definitions of brain-active medications see Supplementary Methods). For each sensitivity analysis, a separate case-control comparison was performed within a subsample retaining only cases with known status for the respective comorbidity variable. Sites that no longer contained both cases and controls following these exclusions were also removed. Within each subsample, two models were estimated per brain region: an unadjusted model (equations 4.2–4.4, excluding β□) and a comorbidity-adjusted model (equations 4.2–4.4, including β□). For each brain region that survived FDR correction in the primary analysis, we assessed whether significance was maintained after adjustment, applying FDR correction to those previously significant regions (see Supplementary Methods for details).

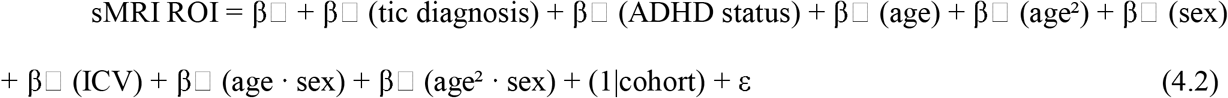

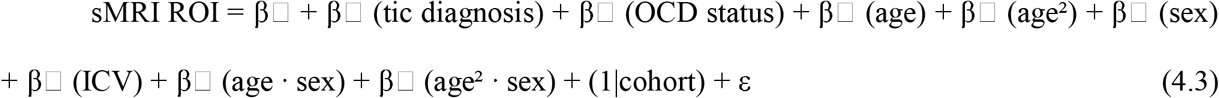

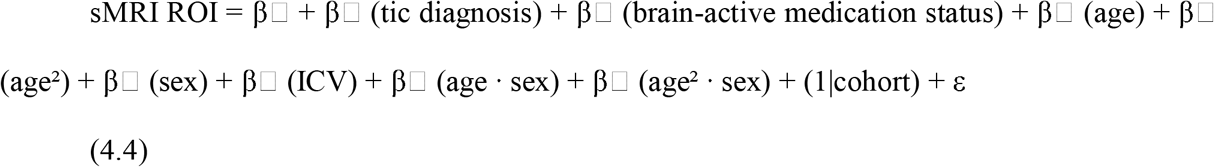

As described in detail above, in our primary analysis, we used linear mixed-effects modeling, widely adopted in multi-site neuroimaging studies ^34,39,43,48^, adjusting for site differences by modeling site-specific means as random intercepts. As additional sensitivity analysis for batch effect, we also applied ComBat-GAM harmonization ^54^, which adjusts for mean and variance across sites via empirical Bayes methods. We defined the dependent, independent, and confounding variables following equation 4.5. Cohen’s *d* ^51^ was derived from the generalized linear model in R. To assess the consistency between the primary mega-analysis and the ComBat-GAM model, Pearson correlations were calculated between effect size estimates.

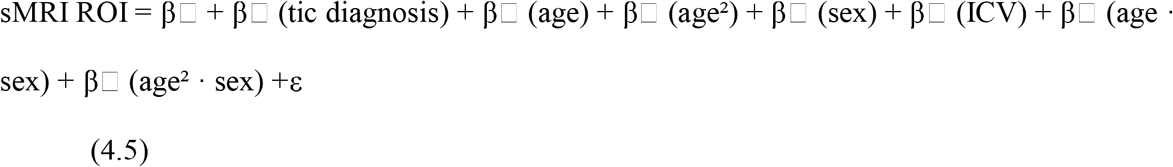

### Brain Effect Comparison between TS/CTD and Other Neuropsychiatric Disorders

To evaluate common and distinct brain mechanisms across TS/CTD and the two disorders most frequently comorbid with TS/CTD (OCD and ADHD), we compared the effect sizes observed in primary mega analysis (Cohen’s *d*) with effect sizes reported in published ENIGMA studies of commonly comorbid disorders. These studies reported effect sizes, compared to healthy controls, for OCD (1,905 cases and 1,760 controls) ^38,41^, ADHD (2,246 cases and 1,934 controls) ^33,34^, with data stratified by children and adults (see Supplementary methods for full details). Notably, for ADHD, analyses were performed on bilaterally averaged ROIs, combining measures from the left and right hemispheres, whereas the OCD study reported left and right ROIs separately. To assess correlations in regional brain effect sizes across disorders while addressing potentially inflated significance from spatial autocorrelation, we applied a spin test ^55^. Specifically, we computed the Pearson correlation between regional effect sizes in TS/CTD and each other disorder and assessed statistical significance using 10,000 spin permutations that preserve spatial structure and control for spatial autocorrelation ^55^. These disorder-level comparisons were conducted separately for adult and pediatric populations.

### Pleiotropy Analysis Between TS/CTD and Brain Structure

To further investigate potential mechanisms linking neuroimaging and genetics, we used summary statistics from the latest TS/CTD genome-wide association study (GWAS), which included 13,247 cases and 536,217 controls ^13^, along with data from the ENIGMA consortium’s GWAS on cortical surface area, thickness, and subcortical volumes ^56,57^. This analysis examined genetic pleiotropy, here meaning that the same single nucleotide polymorphism (SNP) affects both TS/CTD and brain structure, using the SNP Effect Concordance Analysis (SECA) tool ^58^. Detailed methods for SECA are provided in the Supplementary Methods. Since the ENIGMA brain structure GWAS results are not stratified by left and right hemispheres, we reanalyzed our MRI data combining the left and right hemispheres (in the full sample, as well as the pediatric and adult groups separately) to ensure comparability.

## Results

### Sample Characteristics

1,648 participants were included in the analysis following QC, comprising 745 cases (725 with TS and 20 CTDs – here collectively referred to as TS/CTD) and 903 controls scanned in 21 different cohorts (Table 1 and Supplementary Table S2). The dataset includes 510 pediatric cases, 621 pediatric controls, as well as 235 adult cases, and 282 adult controls. The average age at scan was concordant between cases and controls within each age group, with pediatric participants averaging around 11 years old and adults around 31–32 years old (Supplementary Figure S3). The proportion of males was higher in cases compared to controls in both groups. Among patients with known OCD status, comorbid OCD was present in 30% of children and 21% of adults. Similarly, considering TS/CTD cases with known ADHD status, ADHD was reported in approximately 35% of children and 30% of adults. The frequency of OCD and ADHD diagnoses in controls with known comorbidity status was very low across all age groups (0%-14%). For the subset with known brain-active medication status, use was reported in 28% of pediatric cases and 34% of adult cases. More details are provided in Supplementary Table S3.

**Table 1.** Demographic information of TS/CTD patients and controls for 21 cohorts from 13 data providing sites. A total of 1,648 participants with sMRI were analyzed, including 510 pediatric cases, 621 pediatric controls, 235 adult cases, and 282 adult controls. Lifetime diagnoses of ADHD and OCD is reported based on the subset of participants with known OCD and ADHD status

| **Characteristics** | **Pediatric cases** | **Pediatric Control** | **Adult cases** | **Adult Control** |
| --- | --- | --- | --- | --- |
|  | (N = 510) | (N = 621) | (N = 235) | (N = 282) |
| Number of cohorts | 11 | 11 | 10 | 10 |
| **Demographic characteristics** |  |  |  |  |
| Mean age at scan (*mean years ± SD*) | 11 ± 3 | 11 ± 3 | 32 ± 11 | 31 ± 10 |
| Male *N* (%) | 403 (79%) | 391 (63%) | 166 (71%) | 163 (58%) |
| **Clinical characteristics** |  |  |  |  |
| N OCD lifetime diagnosis (N with known OCD status) | 144 (483) | 5 (279) | 26 (123) | 0 (71) |
| N ADHD lifetime diagnosis (N with known ADHD status) | 178 (505) | 49 (343) | 31 (102) | 1 (50) |
| N Brain-active medication uses (N with known brain-active medication status) | 134 (485) | 17 (278) | 49 (146) | 7 (72) |

### Brain Structure Differences in TS/CTD Patients versus Controls from Childhood to Adulthood

We performed a mega-analysis comparing individuals with TS/CTD and controls in the full sample of 745 cases and 903 controls, identifying significant associations between TS/CTD and structural brain measures, including cortical surface area, cortical thickness, and subcortical volumes as detailed below. When considering the full sample across children and adults, TS/CTD was associated with 36 surface area ROIs (global left and right surface area included), 37 cortical thickness ROIs (global left and right thickness included), and four subcortical volumes (Supplementary Table S4 and Figure 1A-C). When considering surface area, the most prominent differences were localized to frontal regions, particularly the orbitofrontal cortex, with the strongest effects observed in the right medial orbitofrontal cortex (FDR-adjusted p = 2.76×10^−10^, Cohen’s d = - 0.378), followed by bilateral lateral orbitofrontal cortex showed significant reductions (right: FDR-adjusted p = 9.08×10^−8^, Cohen’s d = -0.319; left: FDR-adjusted p = 6.21×10^−7^, Cohen’s d = -0.292) (Figure 1A). For cortical thickness, analyses revealed the most prominent reductions in temporal regions, including the middle temporal cortex, temporal pole, and superior and inferior temporal cortices (FDR-adjusted p = 4.67×10^−10^ to 1.57×10^−7^; Cohen’s d = -0.374 to -0.312). In contrast, the most prominent increase was detected in the left pericalcarine cortical thickness (FDR-adjusted p = 2.99×10^−3^; Cohen’s d = 0.179) (Figure 1B). Subcortical volume analyses revealed increased volumes in the bilateral putamen (FDR-adjusted p = 1.13×10^−2^ to 3.29×10^−2^; Cohen’s d = 0.131–0.154) and reduced volumes in the right nucleus accumbens and lateral ventricle (FDR-adjusted p = 4.17×10^−2^and 4.40×10^−2^; Cohen’s d = ™0.125 and −0.120, respectively) (Figure 1C).

**Figure 1.**
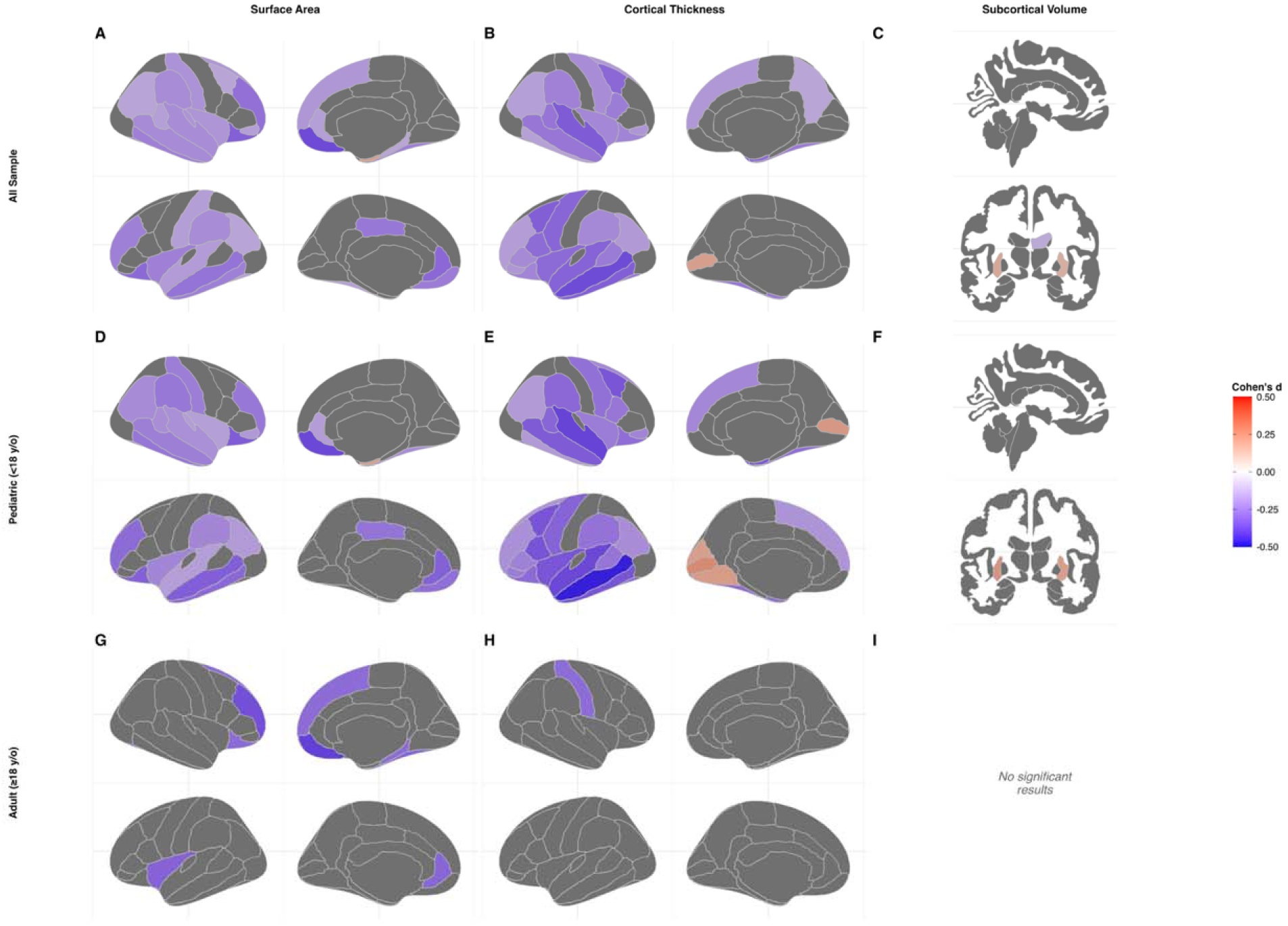
Brain maps illustrating effect sizes (Cohen’s d) of structural differences between individuals with TS/CTD and healthy controls via mega-analysis. Only regions reaching FDR-corrected significance (p < 0.05) are displayed. Non-significant regions are shown in dark grey. Color intensity reflects the magnitude and direction of the effect size: red indicates larger brain volume/thickness/area in TS/CTD relative to controls (positive Cohen’s d), and blue indicates smaller measurements in TS/CTD relative to controls (negative Cohen’s d). Each panel shows cortical maps for the right hemisphere (top row) and left hemisphere (bottom row), with lateral (outer) and medial (inner) views displayed for each hemisphere. (A–C) All ages combined: surface area (A), cortical thickness (B), and subcortical volume (C). (D–F) Pediatric participants (<18 years): surface area (D), cortical thickness (E), and subcortical volume (F). (G–I) Adult participants (≥18 years): surface area (G), cortical thickness (H), and subcortical volume (I). *No significant results* indicates that no brain regions survived FDR correction in that group and measure.

We further performed analysis separately in pediatric and adult groups, seeking to investigate unique brain signatures in each age group. In the pediatric group, TS/CTD was significantly associated with 31 surface area ROIs (global left and right surface area included), 40 cortical thickness ROIs (global left and right thickness included), and three subcortical volumes (Supplementary Table S5 and Figure 1D-F). Among surface area ROIs, the most prominent results were observed for the cortical surface in the right medial orbitofrontal (FDR-adjusted p = 1.13×10^−7^, Cohen’s d = -0.382) and the right lateral orbitofrontal region (FDR-adjusted p = 5.83×10^−6^, Cohen’s d = -0.328) (Figure 1D). The strongest cortical thickness association was thinning observed in the left middle temporal lobe (FDR-adjusted p = 9.93×10^−11^, Cohen’s d = -0.475), followed by the right superior temporal lobe (FDR-adjusted p = 1.13×10^−7^, Cohen’s d = -0.392) and left temporal pole (FDR-adjusted p = 9.58×10^−8^, Cohen’s d = -0.389) (Figure 1E). Increased cortical thickness was observed in occipital regions, including the bilateral pericalcarine cortex, left lingual cortex, and cuneus (FDR-adjusted p = 6.41×10^−4^ to 1.21×10^−2^; Cohen’s d = 0.182 to 0.245). For subcortical volumes, the right nucleus accumbens was significantly smaller (FDR-adjusted p = 6.25×10^−3^, Cohen’s d = -0.200), whereas the left (FDR-adjusted p = 3.37×10^−3^, Cohen’s d = 0.214) and right putamen (FDR-adjusted p = 1.34×10^−2^, Cohen’s d =0.182) had significantly greater volume in the TS/CTD group (Figure 1F).

In adults, eight significant surface area ROIs and two cortical thickness ROIs were identified (Supplementary Table S6 and Figure 1G-I). No subcortical regions showed a significant effect in this dataset which could be related to reduced statistical power due to smaller sample size. Four of the identified surface area measures were in the frontal lobes, including the right lateral orbitofrontal, medial orbitofrontal, rostral middle frontal, and superior frontal regions. Among the surface area regions, the most prominent results were observed in the right medial orbitofrontal (FDR-adjusted p = 6.05×10^−3^, Cohen’s d = -0.403), the right rostral middle frontal (FDR-adjusted p = 1.51×10^−2^, Cohen’s d = -0.363), and the left insula cortex (FDR-adjusted p = 4.63×10^−2^, Cohen’s d = -0.314) (Figure 1G). The cortical thickness analysis showed significantly lower cortical thickness in the right temporal pole cortex and right postcentral cortex (both FDR-adjusted p = 4.63×10^−2^, Cohen’s d = - 0.289) (Figure 1H).

Meta-analyses were conducted in the full sample, as well as in pediatric and adult subgroups (see Supplementary Results, Supplementary Figure S4, and Supplementary Tables S7–S9). Pearson correlation analyses demonstrated high agreement between the meta-analysis and the primary mega-analysis, with correlations of r = 0.89 in the full sample, r = 0.90 in children, and r = 0.93 in adults.

### Age- and Sex-by-Diagnosis Effects on Brain Structure in TS/CTD

To assess the effects of age and sex on TS/CTD case-control differences in brain structure, we tested age-by-diagnosis (including both linear and quadratic age terms) and sex-by-diagnosis interactions for each ROI across the full sample and each age group. In the full sample, significant quadratic age-by-diagnosis (age^2^) interactions were observed in cortical thickness analysis, involving 22 ROIs (all FDR < 0.05). Furthermore, in the pediatric-only group, significant quadratic age-by-diagnosis (age^2^) interactions were observed, specifically in the bilateral thalamus (right thalamus: β = –3.487 × 10^3^, FDR-adjusted p = 2.42×10^−2^; left thalamus: β = −3.461×10^3^, FDR-adjusted p = 2.58×10^−2^; Supplementary Figure S5). No significant linear age-by-diagnosis or sex-by-diagnosis interactions were observed in the full sample or within the pediatric and adult groups, and no significant quadratic age-by-diagnosis interactions were detected in the adult group (Supplementary Tables S10-S12).

### Sensitivity analyses

To assess the robustness of the primary findings to comorbid ADHD, OCD, and brain-active medication use, sensitivity analyses were conducted separately in subsamples with available information for each condition. Overall, the majority of significant brain structural differences identified in the unadjusted analysis were maintained after additional adjustment for these factors and despite the smaller sample available for analysis with known comorbidity status. For ADHD, 66 of 73 regions remained significant in the overall subsample (90.4%; 607 cases, 815 controls), 67 of 74 in children (90.5%; 505 cases, 621 controls), and 16 of 20 in adults (80.0%; 102 cases, 194 controls). For OCD, 69 of 71 regions remained significant in the overall subsample (97.2%; 606 cases, 773 controls), 70 of 72 in children (97.2%; 483 cases, 557 controls), and 5 of 5 in adults (100%; 123 cases, 197 controls). For brain-active medication use, 69 of 72 regions remained significant in the overall subsample (95.8%; 631 cases, 773 controls), 70 of 73 in children (95.9%; 485 cases, 557 controls), and 13 of 13 in adults (100%; 146 cases, 216 controls). Overall, robustness was generally high across all our sensitivity analyses, with near-complete retention when accounting for OCD and brain-active medication use and slightly lower retention in the ADHD-adjusted analysis in adults. This likely reflects reduced statistical power in smaller adult subsamples. Full results are reported in Supplementary Tables S13–S21.

Furthermore, although our primary analysis accounted for batch effects using a mixed-effects model, we additionally applied ComBat-GAM harmonization as an additional/alternative approach for multi-site comparisons. In the full sample, 71 significant ROIs were identified, all of which were also detected in the primary mega-analysis (Supplementary Table S22). In the pediatric sample, this analysis identified 74 significant ROIs, 73 (98.6%) of which overlapped with the primary mega-analysis (Supplementary Table S23). In adults, where the sample size was substantially smaller, ComBat-GAM analysis resulted in reduced statistical power and yielded no significant findings (Supplementary Table S24). Correlation analyses comparing the ComBat-GAM harmonization approach with the primary mixed-effects model showed very high agreement across the full, pediatric, and adult samples (r = 0.97–0.99), supporting the overall consistency of regional effect patterns across methods (Supplementary Table S25).

### Cross-Disorder Comparison of Brain Structure in TS/CTD

We compared our TS/CTD sMRI results with ENIGMA sMRI studies in OCD and ADHD (the two most common comorbid conditions in TS/CTD) to investigate shared and distinct brain alterations in each age group (Figure 2, Supplementary Table S26). In the pediatric group, TS/CTD effects on cortical surface area were significantly positively correlated with those of ADHD (r = 0.57, p_perm_ = 5 × 10^−4^; Figure 2A) and OCD (r = 0.35, p_perm_ = 1.0 × 10^−3^; Figure 2C). Additionally, a positive correlation was observed between TS/CTD and ADHD in their effects on cortical thickness (r = 0.37, p_perm_ = 1.5×10^−3^; Figure 2A). In the adult group, the brain effect of TS/CTD had a significant positive correlation with ADHD in surface area (r = 0.27, p_perm_ = 1.90×10^−2^; Figure 2B) and subcortical regions (r = 0.34, p_perm_ = 2.00×10^−2^; Figure 2B). The effects of TS/CTD on the brain were also correlated with OCD subcortical volumes in adults (r = 0.60, p_perm_ = 1.0×10^−2^; Figure 2D).

**Figure 2.**
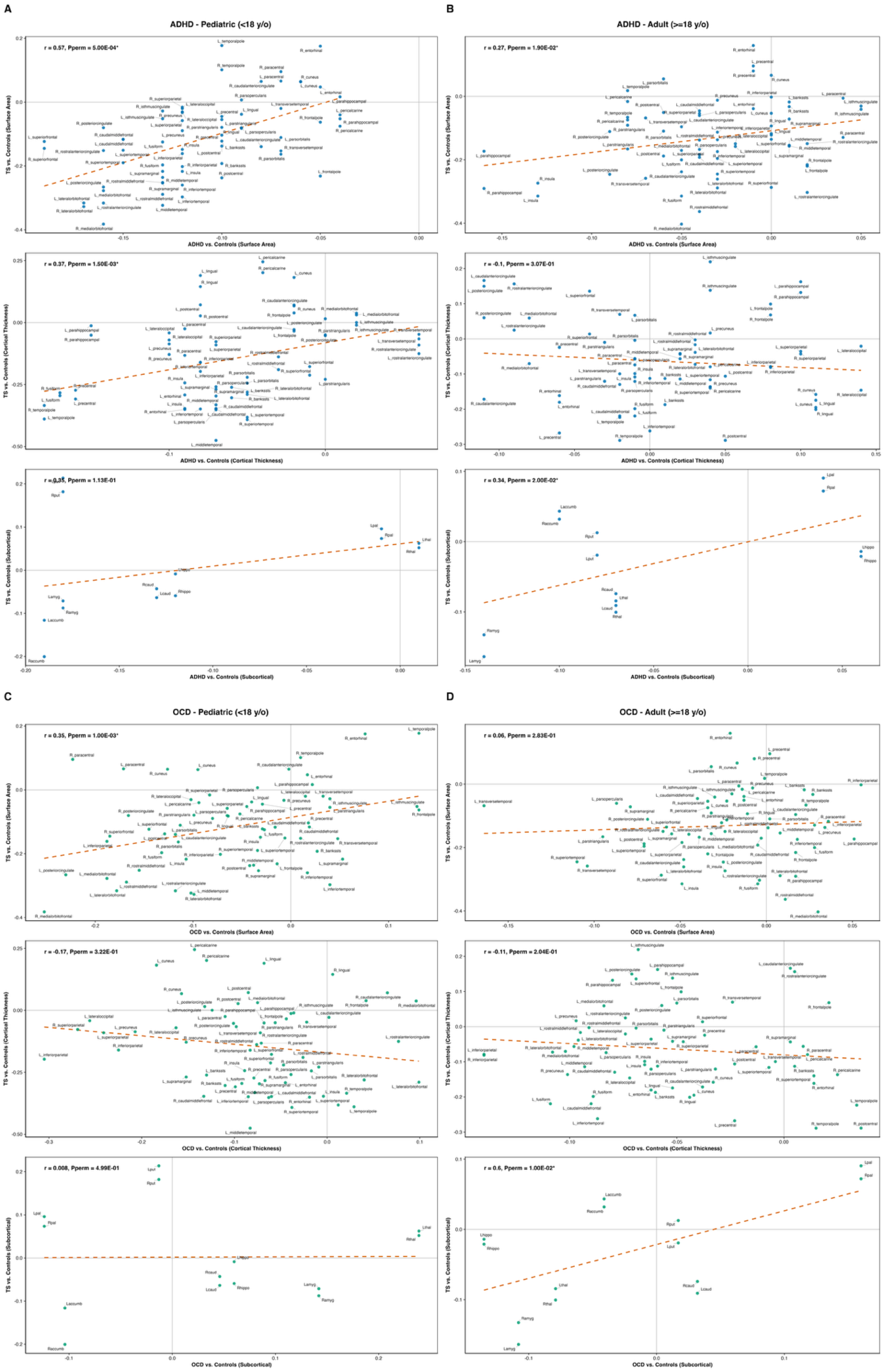
Cross-disorder comparison of brain structure in TS/CTD and frequently comorbid psychiatric disorders. Scatter plots illustrating correlations of regional brain effect sizes (Cohen’s d) between TS/CTD and its two most common comorbid disorders (ADHD and OCD). Each point represents an individual brain region. The x-axis shows Cohen’s d for the respective comorbid disorder versus controls, and the y-axis shows Cohen’s d for TS/CTD versus controls. The dashed orange line indicates the linear regression fit. Pearson correlation coefficients (r) and spin test permutation P value are displayed in the upper-left corner of each panel; asterisks (*) denote statistically significant correlations (Pperm* < 0.05). Brain regions are labeled with anatomical names. (A) ADHD Pediatric (<18 years): surface area (top), cortical thickness (middle), and subcortical volume (bottom). (B) ADHD Adult (≥18 years): surface area (top), cortical thickness (middle), and subcortical volume (bottom). (C) OCD Pediatric (<18 years): surface area (top), cortical thickness (middle), and subcortical volume (bottom). (C) OCD Adult (≥18 years): surface area (top), cortical thickness (middle), and subcortical volume (bottom). Blue points correspond to ADHD (A–B) and teal points to OCD (C–D).

### Genetic Pleiotropy between TS/CTD risk and Brain Structure

We pursued genetic pleiotropy analyses to test whether loci associated with TS/CTD risk also influence brain structure, as identified in large-scale GWAS (see Methods). Indeed, we found significant global pleiotropy between TS/CTD genetic risk variants and variants associated with 11 cortical surface area regions, four cortical thickness regions, and four subcortical volumes (FDR-corrected; Supplementary Table S27 and Figure 3A–B).

**Figure 3.**
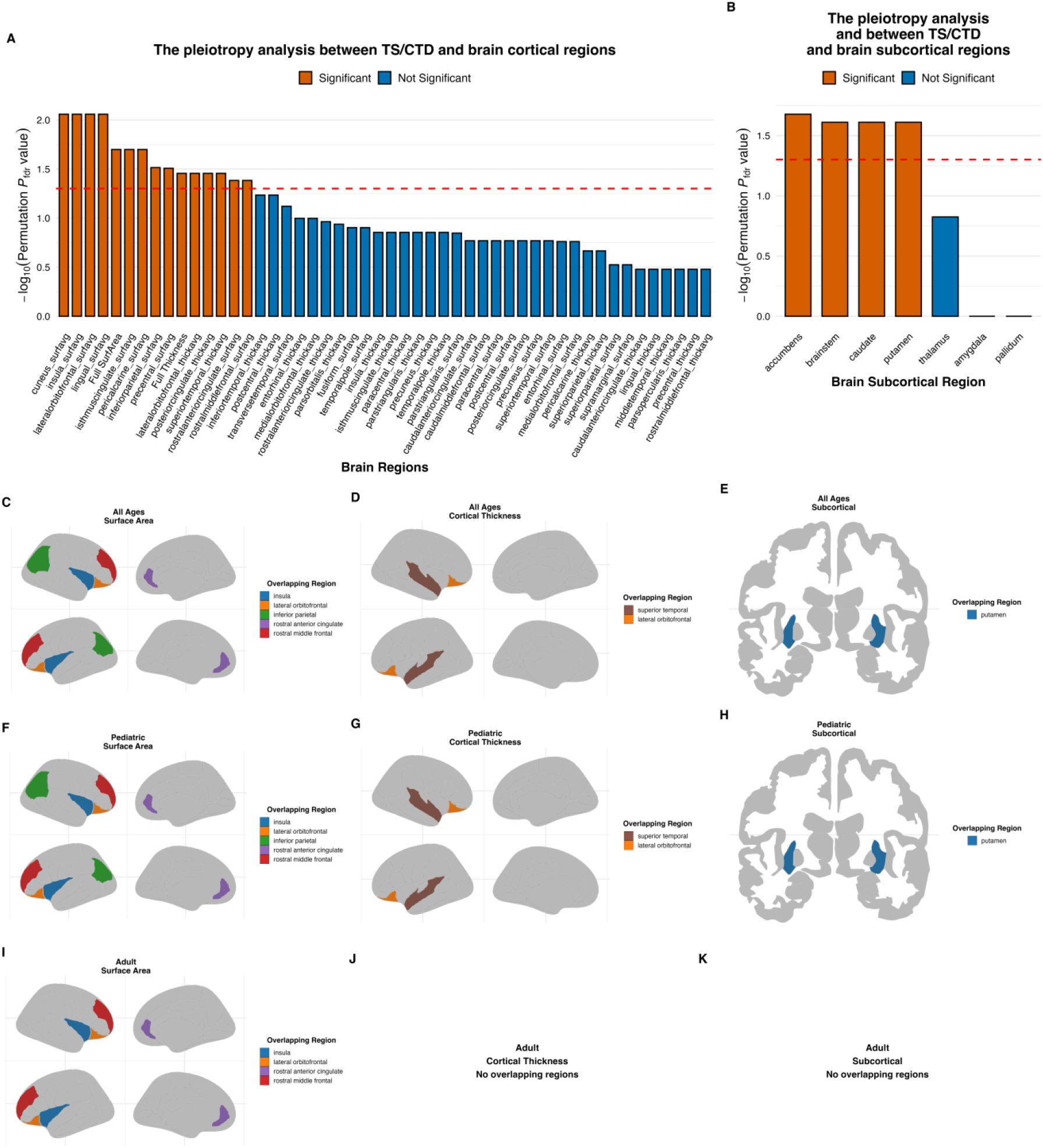
Pleiotropic genetic overlap and structural brain alterations in TS/CTD. (A) Bar plot showing –log_10_-transformed FDR-corrected permutation P values for pleiotropic genetic overlap between TS/CTD and cortical brain regions derived from GWAS, estimated using the SNP Effect Concordance Analysis (SECA) tool. The dashed red line indicates the significance threshold (FDR P < 0.05). Orange bars denote regions reaching statistical significance; blue bars denote non-significant regions. (B) Bar plot showing –log_10_-transformed FDR-corrected permutation P values for pleiotropic genetic overlap between TS/CTD and subcortical brain regions. The dashed red line indicates the significance threshold (FDR P < 0.05); color coding is as in panel A. (C–K) Brain maps depicting regions with significant structural alterations in TS/CTD from the ENIGMA mega-analysis that overlap with regions showing significant pleiotropic genetic effects, shown separately for surface area, cortical thickness, and subcortical volume, and stratified by age group. Regions without overlap are shown in grey. Each panel shows cortical maps for the right hemisphere (top row) and left hemisphere (bottom row), with lateral (outer) and medial (inner) views displayed for each hemisphere. (C–E) All participants: surface area (C), cortical thickness (D), and subcortical volume (E). (F–H) Pediatric participants (<18 years): surface area (F), cortical thickness (G), and subcortical volume (H). (I–K) Adult participants (≥18 years): surface area (I), cortical thickness (J), and subcortical volume (K). **Note:** The nucleus accumbens could not be visualized.

The brain regions identified through pleiotropy analyses as sharing genetic drivers with TS/CTD risk were compared with findings from our neuroimaging mega-analysis of bilaterally averaged measures to evaluate alignment with case–control structural differences (Supplementary Tables S28–S30). In both the full sample and the pediatric group, six surface area regions (including global surface area) and three cortical thickness regions (including global cortical thickness) showed both significant pleiotropic overlap with TS/CTD GWAS and structural alterations associated with TS/CTD (Figure 3C–D, F-G). Among subcortical structures, the putamen showed both pleiotropic genetic overlap with TS/CTD risk and significant case–control differences in the pediatric and full samples, whereas the nucleus accumbens showed this pattern in the pediatric group only (Figure 3E and 3H). In adults, four surface area measures with significant pleiotropic overlap between TS/CTD GWAS and brain structure GWAS also showed corresponding structural differences in neuroimaging analyses (Figure 3I). Notably, several surface area ROIs consistently associated with TS/CTD across neuroimaging analyses in all studied groups —including the lateral orbitofrontal cortex, rostral middle frontal cortex, insula, and rostral anterior cingulate cortex—also showed evidence of genetic pleiotropy with TS/CTD. Together, these findings demonstrate convergence between genetic risk for TS/CTD, and neuroanatomical alterations observed in patients.

## Discussion

This study provides the most comprehensive characterization to date of how brain structure is altered across development in TS/CTD, integrating (post-QC) data from 1,648 participants across 21 international cohorts within the ENIGMA-TS consortium. Our results reinforce the central role of CSTC circuitry in TS/CTD while also indicating involvement of additional frontal and paralimbic regions such as the insula and anterior cingulate. These findings suggest that alterations in CSTC circuits may interact with broader cortical systems involved in cognitive control, salience processing, and sensory integration, and support the view that TS/CTD reflects not simply a motor disorder, but a disorder of inhibitory control and urge regulation involving frontal–striatal networks.

Studying brain structure from childhood to adulthood, we observed widespread structural alterations, with all significant age-by-diagnosis interaction effects localized to cortical thickness. As cortical thickness is strongly influenced by neurodevelopmental processes that occur later in life, including synaptic pruning and myelination ^59,60^, these results reframe TS/CTD as disorders that are associated with altered trajectories of cortical maturation and potential compensatory processes rather than static structural differences. Stratifying analyses by developmental stage, we revealed different patterns in pediatric and adult participants. Unlike the more circumscribed profiles identified for other psychiatric disorders in ENIGMA, such as ADHD, and OCD ^33,34,38,41^, which emphasized select frontal and subcortical loci, alterations in pediatric TS/CTD were broader and more diffuse. On the other hand, in adults, effects were more focal and largely restricted to frontal regions.

Among the significant regions in children, those showing the strongest statistical effects were key components of the multiple CSTC circuits (associative, sensorimotor, and cognitive), including prefrontal and orbitofrontal cortices, sensorimotor regions, the anterior cingulate cortex, and the basal ganglia ^32,61–63^. Notably, the orbitofrontal cortex, a region central to behavioral inhibition, habit learning, and emotional regulation, showed consistently lower surface area and cortical thickness ^20,22^. In addition, the putamen, a striatal region essential for motor control and procedural learning ^64,65^, demonstrated greater volume in TS/CTD in our analysis of the pediatric cohort. Beyond frontostriatal alterations, significant effects were also observed in sensorimotor cortices, including the primary motor cortex, primary somatosensory cortex, supplementary motor area, and cingulate motor cortex. Prior studies have likewise reported sensorimotor cortical thinning in TS/CTD, implicating these regions in tic generation ^66,67^. These findings further support the CSTC model in TS/CTD, indicating that dysregulated signaling across motor, associative, and regulatory circuits contributes to tic generation and modulation.

However, structural alterations in pediatric TS/CTD were not confined to the CSTC circuit. For example, significant effects were also observed in lateral temporal and parietal association cortices, paralimbic regions such as the entorhinal cortex and insula, as well as occipital cortex. Lateral temporal and parietal association cortices are integral to attention to stimuli, with fMRI studies in TS/CTD reporting engagement of the temporoparietal junction during mental state reasoning tasks ^68^. In addition, paralimbic structures, particularly the entorhinal cortex, exhibited some of the largest effect sizes in cortical thickness in the meta-analysis, consistent with previous reports linking entorhinal abnormalities to symptom modulation ^32^. In contrast to cortical thinning observed in other regions, several regions in occipital cortex showed increased thickness relative to controls. This pattern may reflect heightened sensitivity to visual stimuli, which have been reported as potential triggers for tic expression ^69^. This widespread structural involvement suggests that TS/CTD reflects a distributed neurodevelopmental condition, potentially affecting multiple interacting functional systems rather than a single isolated pathway.

In contrast, adult TS/CTD participants exhibited more focal effects, restricted primarily to surface area alterations within the orbitofrontal and rostral middle frontal cortices. This developmental divergence is consistent with the typical clinical trajectory of TS/CTD in which symptoms often improve during adolescence and early adulthood ^1^.

Our finding of significant quadratic age-by-diagnosis (age^2^) interactions in bilateral thalamic volume among the pediatric TS/CTD group may account for the mixed findings reported in earlier studies, which described both greater and lower thalamic volumes, or no significant differences at all ^18,19,26,70^, often without modeling age effects in sufficient detail. The nonlinear pattern indicates that case–control differences vary across development during childhood. Our results thus point to the thalamus as a dynamic node in the CSTC network whose role likely shifts with maturation, consistent with observations in OCD ^71^. Given the integrative function of the thalamus in sensory and motor gating, its evolving structural profile may reflect both the progression of tic pathology and the brain’s compensatory responses during adolescence, a period marked by synaptic reorganization and cortical–subcortical recalibration.

Genetic pleiotropy analyses indicated shared genetic influences between TS/CTD and specific cortical and subcortical structures several of which were found altered in patients versus controls in our neuroimaging analyses. It is important to note that pleiotropy, in which genetic variants can affect multiple phenotypes, indicates association but does not necessarily show a direct causal relationship. Among brain regions significantly associated with TS/CTD across all our analyses, genes influencing the lateral orbitofrontal cortex, rostral middle frontal cortex, insula, and rostral anterior cingulate cortex also showed pleiotropy with TS/CTD genetic risk. In pediatric analyses specifically, pleiotropic overlap also included subcortical structures such as the putamen and nucleus accumbens, aligning with previous findings linking striatal structures to tic disorders ^72^. Frontal cortical involvement further supports a genetic basis for CSTC dysregulation. The insula, implicated in limbic motor hyperactivation associated with tics and the urge to tic ^73,74^, and the rostral anterior cingulate cortex, related to tic severity and urge intensity possibly reflecting reduced GABAergic inhibition ^22,75,76^, underscore potential genetic pathways relevant to tic expression. A previous fMRI study also found increased connectivity in TS between the insula, putamen, orbitofrontal, and cingulate cortices ^77^.

Further contextualizing these structural brain alterations within established functional networks provides additional insights. The insula, the putamen and the thalamus figure prominently in the recently renamed action mode network (AMN), involved in decision-making and action initiation ^78,79^. Conversely, the orbitofrontal and entorhinal cortices belong to the default mode network (DMN) ^80^, while the middle frontal gyrus participates in executive control via the frontoparietal network. The putamen’s functional connectivity shifts from facial somatomotor networks in children to control and attention networks in adults ^81^. AMN dysfunction has been implicated in tic generation ^82^, supported by lesion studies linking insula, putamen, and thalamus damage to tics, and negatively associating the precuneus (DMN) with tic causation ^83^. Insula and thalamus involvement in various urges ^76^ and associations between insula thinning and urge-to-tic further underscore their importance ^25^. These findings collectively support a model in which disrupted balance between internally focused (DMN), and action-oriented (AMN) states contributes to TS/CTD, with structural alterations reflecting or driving this imbalance. Consistent with this interpretation, recent fMRI studies in TS have reported widespread functional alterations across multiple brain networks, ^84^ for instance, somatomotor networks, DMN, frontoparietal control, and attention networks. The breadth of these functional abnormalities aligns with the distributed structural alterations observed here.

Integration with data from the two most frequently comorbid psychiatric disorders in TS/CTD highlighted significant cross-disorder correlations and once again, highlighted different patterns across development. TS/CTD -related brain structural effects showed significant positive spatial correlations with ADHD and OCD across age groups, with a clear developmental dissociation: cortical correlations predominated in children, whereas subcortical correlations were stronger in adults. These cross-disorder patterns are consistent with partially shared neurobiological foundations, particularly within cortico-subcortical loops that govern motor control, cognitive flexibility, and emotional regulation ^17,31–34,36,37,85^. The observed convergence with ADHD and OCD is consistent with epidemiological comorbidity and overlapping CSTC involvement. A transdiagnostic approach that could leverage pooled individual-level neuroimaging data ^35^ could further delineate shared and disorder-specific neuroanatomical profiles and represents an important direction for future work. Future studies can also include comparisons with additional disorders that are correlated or comorbid with TS/CTD to yield further insights.

Several limitations should be noted. Although the present study leverages unprecedented sample size and harmonized imaging processing and analysis protocols, its cross-sectional design limits inference about developmental trajectories. Furthermore, this design cannot determine whether the structural brain differences in TS/CTD constitute a cause or a consequence of having tics. Future longitudinal and multimodal imaging will be essential to directly capture neurodevelopmental change and its relation to symptom progression. Medication use was classified only as brain-active versus non–brain-active in this study; future analyses with larger sample sizes will more precisely assess medication-specific effects on brain structure in TS/CTD. The subsamples used in the sensitivity analyses require larger sample sizes to reduce the risk of false-positive findings and improve the stability of effect estimates. Variability in imaging acquisition, clinical assessments and comorbidity measures across sites may have introduced residual heterogeneity, and the pleiotropy analyses were constrained by current GWAS sample sizes for TS/CTD, although the largest available datasets were used. Incorporating functional, diffusion, and transcriptomic data will further refine the mechanistic links between genetic variation and neural circuitry.

Taken together, our findings delineate a distributed and developmentally dynamic neuroarchitecture of TS/CTD that is shaped by genetic influences and evolves across development.

In pediatric populations, TS/CTD manifests as coordinated and widespread alterations across both cortical and subcortical regions, extending beyond canonical CSTC circuitry, whereas in adults these alterations are more focal and predominantly localized to frontal systems. This pattern supports a model in which TS/CTD arise from altered neurodevelopmental trajectories that evolve across the lifespan and may partially normalize through maturation and compensatory processes. The large-scale, harmonized nature of the ENIGMA-TS dataset, the integration with genomic studies, and the transdiagnostic context establish a strong foundation for future work aimed at biomarker discovery, elucidation of causal mechanisms, and the development of targeted, developmentally informed interventions.

## Data Availability

All data produced in the present study are available upon reasonable request to the authors

## Consortia

**ENIGMA-TS Consortium:** Jan Buitelaar, Valerie Brandt, Francesco Cardona, Danielle Cath, Giulia Conte, Catharina Cramer, Nanette Mol Debes, Heike Eichele, Natalie Forde, Barbara Franke, Pablo Franco-Rosado, Christos Ganos, Deanna J. Greene, Michel Grothe, Yuxin Guo, Andreas Hartmann, Theresa Valentine Heinen, Tamara Hershey, Neda Jahanshad, Piotr Janik, Yin Jin, Jon Koller, Anne Kleine Büning, Kirsten Mueller-Vahl, Karsten Mueller, Alexander Munchau, Richard Musil, Juan Francisco Martin-Rodriguez, Pablo Mir, Harald Möller, Edoardo Monfrini, Irene Neuner, Stefano Palmucci, Mauro Porta, Adriana Prato, Shukti Ramkiran, Renata Rizzo, Veit Roessner, Federica Saia, Katharina Anna Schindlbeck, Bradley L. Schlaggar, Tim J. Silk, Natalia Szejko, Sophia Thomopoulos, Paul M. Thompson, Silvia Tommasin, Apostolia Topaloudi, Renzo Torrecuso, Zeynep Tumer, Anne Uhlmann, Odile van den Heuvel, Dick Veltman, Julius Verrel, Tanja Veselinovic, Yulia Worbe, Tomasz Wolanczyk, Laura Zapparoli, Jade-Jocelyne Zouki, Kevin J. Black, Peristera Paschou

**TAA Neuroimaging Consortium:** Adriana Di Martino, F. Xavier Castellanos, Barbara J. Coffey, Michael P. Milham, Abigail Mengers, Krishna Somandepalli, Stewart H. Mostofsky, Harvey S. Singer, Carrie Nettles, Daniel Peterson, Deanna Crocetti, John Piacentini, Susan Y. Bookheimer, James T. McCracken, Susanna Chang, Adriana Galvan, Kevin J. Terashima, Elizabeth R. Sowell, Bradley L. Schlaggar, Kevin J. Black, Deanna J. Greene, Jessica A. Church, Steven E. Petersen, Tamara Hershey, Deanna M. Barch, Joan L. Luby, Alton C. Williams, Jonathan M. Koller, Matthew T. Perry, Emily C. Bihun, Samantha A. Ranck

## TAAICG

Dongmei Yu, Zachary Gerring, Tyne W. Miller-Fleming, Luz M. Porras, Franjo Ivankovic, Cathy L. Barr, James R. Batterson, Fortu Benarroch, Cathy L. Budman, Danielle Cath, Laura Domènech, Emily Gantz, Marco A. Grados, Erica L. Greenberg, Luis Diego Herrera-Amighetti, Alden Y. Huang, David Isaacs, Joseph Jankovic, James F. Leckman, Christine Lochner, Irene A. Malaty, William M. McMahon, Benjamin M. Neale, Erika Nurmi, Michael S. Okun, David L. Pauls, Danielle Posthuma, Mary Robertson, Guy Rouleau, Paul Sandor, Joshua A. Senior, Harvey S. Singer, Lea K. Davis, Paola Giusti-Rodríguez, Peristera Paschou, Jeremiah M Scharf, Carol A Mathews

**EMTICS:** Apostolia Topaloudi, Sudhanshu Shekhar, Alan Apter, Valentina Baglioni, Juliane Ball, Noa Benaroya-Milshtein, Judith Buse, Francesco Cardona, Marta Correa Vela, Andrea Dietrich, Blanca Garcia-Delgar, Julie Hagstrøm, Tammy J. Hedderly, Isobel Heyman, Pieter J. Hoekstra, Chaim Huijser, Marcos Madruga-Garrido, Anna Marotta, Pablo Mir, Astrid Morer, Norbert Müller, Kirsten Müller-Vahl, Alexander Münchau, Peter Nagy, Kerstin J. Plessen, Cesare Porcelli, Renata Rizzo, Veit Roessner, Tamar Steinberg, Zsanett Tarnok, Susanne Walitza, Elif Weidinger, Jin Yin, Peristera Paschou

**TS-EUROTRAIN/TSGeneSEE:** Christos Androutsos, Csaba Barta, Entela Basha, Dorret I. Boomsma, Jan K. Buitelaar, Christel Depienne, Andrea Dietrich, Petros Drineas, Gudmundur Einarsson, Siyan Fan, Jakub P Fichna, Natalie J. Forde, Abel Fothi, Marianthi Georgitsi, Jeffrey Glennon, Daniel F. Gudbjartsson, Andreas Hartmann, Bastian Hengerer, Pieter J. Hoekstra, Piotr Janik, Cathrine Jespersgaard, Ahmad Seif Kanaan, Mira Kapisyzi, Iordanis Karagiannidis, Anastasia Koumoula, Shanmukha S Padmanabhuni, Geert Poelmans, Petra J. W. Pouwels, Joanna Puchala, Natalia Szejko, Urszula Szymanska, Olafur Thorarensen, Zeynep Tümer, Odile A. van den Heuvel, Ysbrand D. van der Werf, Dick J. Veltman, G. Bragi Walters, Joanna Widomska, Tomasz Wolanczyk, Yulia Worbe, Cezary Zekanowski, Nuno R. Zilhäo, Hreinn Stefansson, Peristera Paschou

## TIC Genetics

The members of the Tourette International Collaborative Genetics (TIC Genetics) consortium (in alphabetical order): Juliane Ball, Noa Benaroya-Milshtein, Kate Bornais, Keun-Ah Cheon, Barbara J. Coffey, Andrea Dietrich, Erik M. Elster, Dana Feldman, Thomas V. Fernandez, Carolin Fremer, Danielle Cath, Donald L. Gilbert, Danea Glover, Tammy Hedderly, Gary A. Heiman, Isobel Heyman, Pieter J. Hoekstra, Hyun Ju Hong, Chaim Huijser, Christina Kappler-Friedrichs, Young Key Kim, Young Shin Kim, Robert A. King, Nadine Kirchen, Carolin Sophie Klages, Samuel Kuperman, Bennett L. Leventhal, Holan Liang, Maria Loreta Lopez, Osman Malik, Marieke Messchendorp, Dararat Mingbunjerdsuk, Pablo Mir, Astrid Morer, Kirsten R. Müller-Vahl, Alexander Münchau, Laura Muñoz-Delgado, Tara L. Murphy, Cara Nasello, Kerstin J. Plessen, Veit Roessner, Alyssa Rosen, Guy Rouleau, Simon Schmitt, Chitra Shukla, Sara Sopena, Matthew W. State, Tamar Steinberg, Zsanett Tarnok, Joshua K. Thackray, Meitar Timmor, Jay A. Tischfield, Max A. Tischfield, Anne Uhlmann, Ana Vigil-Pérez, Frank Visscher, Susanne Walitza, Belinda Wang, Sheng Wang, A. Jeremy Willsey, Jinchuan Xing, Samuel H. Zinner.

NORDIC Sweden: David Mataix-Cols, Christian Rück, Julia Bäckman, Elles de Schipper, Per Andrén, James Crowley, Matthew W Halvorsen

