## Supplementary Figures and Methods for "Altered neurodevelopmental trajectories of brain structure in Tourette syndrome and Chronic Tic Disorders"

**”**

[References	1](#_Toc229393039)3

### Supplementary Methods

#### Brain-active medications

Brain-active medications were defined as pharmacological agents with direct central nervous system effects and were categorized into the following classes: selective serotonin reuptake inhibitors (SSRIs), α2-adrenergic agonists, dopamine antagonists, stimulants, benzodiazepines, other antidepressants, norepinephrine (NE) reuptake inhibitors, vesicular monoamine transporter 2 (VMAT2) inhibitors, and other brain-active medications not captured by these categories. All remaining medications without known primary central nervous system effects were classified as non–brain-active.

#### Run FreeSurfer and Cortical & Subcortical Measures Extraction

The ENIGMA FreeSurfer protocol (<https://enigma.ini.usc.edu/protocols/imaging-protocols/>) establishes a harmonized framework for processing, extracting, and quality-controlling structural MRI data across international cohorts. The workflow begins by processing each subject’s T1-weighted MRI scan through FreeSurfer’s recon-all command, which performs automated cortical reconstruction and volumetric segmentation. This process includes skull stripping, intensity normalization, registration to a standardized space, segmentation of subcortical structures, and the generation of cortical surface models. It further performs surface inflation and cortical parcellation based on the Desikan–Killiany atlas ^1^, generating measures of cortical thickness and surface area, along with ASEG atlas ^2^ estimates of subcortical volumes. By standardizing this key preprocessing step, ENIGMA ensures that data derived from different scanners, protocols, and populations are directly comparable. Following reconstruction, ENIGMA’s wrapper scripts automate the extraction of regional statistics. The pipeline generates summary CSV files for cortical thickness, surface area, and subcortical volumes.

#### Quality control (QC) pipeline

Based on the ENIGMA quality control protocols (https://enigma.ini.usc.edu/protocols/imaging-protocols/), a comprehensive two-stage QC procedure was implemented for both cortical and subcortical measures derived from FreeSurfer processing. For cortical data, the process began with outlier detection using an R script to identify subjects with extreme cortical thickness and surface area values. This was followed by visual inspection using two methods: the internal surface method generated cross-sectional images of segmentations overlaid on anatomical scans via a custom MATLAB script, while the external surface method produced lateral and medial views of the pial surface using a shell script.

For subcortical volumes, a parallel QC workflow was executed, commencing with the generation of histogram plots to visualize the distribution of volumetric data for each structure. Automated outlier detection was then performed using a shell script to flag subjects with volumes falling outside the expected interquartile interval. Flagged subjects underwent direct visual inspection in FSLview, where subcortical segmentations were overlaid on native T1 scans. Finally, a series of standardized PNG snapshots were generated for all subjects using a MATLAB script and compiled into a subcortical-specific QC webpage. All visual outputs were compiled into HTML webpages for systematic rating against the ENIGMA Cortical and Subcortical Control Guide.

Following ENIGMA-established visual quality control procedures, 52 scans were excluded due to insufficient image quality, yielding 2,092 scans from 1,756 participants. Scans with missing covariate information were subsequently removed, including 30 lacking age and/or sex, resulting in 2,062 scans from 1,726 participants. Individuals with provisional or transient tic disorders (PTD/TTD; scans = 87) were excluded, leaving 1,975 scans from 1,700 participants. One participant receiving trientine treatment, indicative of Wilson’s disease, was removed due to clinical incompatibility with the study aims, yielding 1,974 scans from 1,699 participants. To ensure independence of observations, three related individuals (twins/siblings) were excluded, resulting in 1,971 scans from 1,696 participants. For individuals with multiple scans, a single scan per participant was retained, removing 275 repeated scans and yielding a cross-sectional dataset of 1,696 scans from 1,696 participants. An additional 47 participants were excluded due to insufficient sample size within strata defined by age group and scanner characteristics, leaving 1,649 scans. Finally, one participant identified as an intracranial volume (ICV) outlier based on Cook’s distance was excluded. The final analytic sample comprised 1,648 scans from 1,648 individuals. The complete QC workflow is presented in Supplementary Figure S2.

#### Comparison of Brain Differences Between TS/CTD and Controls: A Meta-Analysis

To maintain consistency with prior ENIGMA studies ^3–5^ and leverage the advantages of multisite neuroimaging, we also conducted a meta-analysis. This allowed for direct comparisons across different research cohorts. The dependent, independent, and confounding variables were aligned with those used in the primary mega-analysis (Supplementary equation S1.1). Regression models and effect size estimates were calculated separately for each cohort to account for site-specific variability. Cohen's *d* ^6^ was derived from the generalized linear model in R. A false discovery rate threshold of < 0.05 was applied as the criterion for statistical significance. To assess the consistency between the primary mega- and meta-analyses, we conducted Pearson correlation tests.

sMRI ROI = β₀ + β₁ * (tic diagnosis) + β₂ * (age) + β₃ * (age²) + β₄ * (sex) + β₅ * (ICV) + β₆ * (age * sex) + β₇ * (age² * sex) + ε	(S1.1)

#### Statistical Power Analysis

To evaluate the statistical adequacy of the ENIGMA-TS sample, we conducted a post hoc power analysis using the pwr package (v1.3-0)^7^ in R. Given the multi-site design, power was estimated using a two-sample t-test approximation, which is conservative relative to the linear mixed-effects models used in the primary analyses. Power was calculated separately for the full sample (745 cases, 903 controls; N = 1,648), the pediatric subgroup (510 cases, 621 controls; N = 1,131), and the adult subgroup (235 cases, 282 controls; N = 517), at a two-sided significance level of α = 0.05, consistent with the false discovery rate threshold applied in the primary analyses. At 80% power, the minimum detectable effect sizes were Cohen’s d = 0.139 for the full sample, d = 0.168 for the pediatric subgroup, and d = 0.248 for the adult subgroup. Of the 77 FDR-significant regions in the full sample (Cohen’s d = 0.120–0.378), 68 regions (88.3%) exceeded the 80% power threshold, with most remaining regions exceeding 70% power. Similarly, of the 74 FDR-significant regions in the pediatric subgroup (Cohen’s d = 0.148–0.475), 66 regions (89.2%) exceeded 80% power, with most remaining regions also above 70% power. In contrast, the adult subgroup achieved 90.0% power at its smallest significant effect (Cohen’s d = 0.287), with all 10 FDR-significant regions (Cohen’s d = 0.287–0.403) robustly exceeding the 80% detection threshold.

#### Sensitivity analysis

A series of sensitivity analyses was conducted to examine whether brain structural differences associated with TS/CTD are robust to the confounding effects of comorbid ADHD, OCD, and brain-active medication use. For each sensitivity analysis, a separate case-control comparison was performed within a subsample retaining only cases with known status for the respective comorbidity variable. All TS/CTD cases with known comorbidity status were retained in each analysis, with comorbidity status included as a binary covariate in the model (1 = present/active, 0 = absent). For controls, comorbidity status was assigned based on known status where available and coded as 0 when status was unknown. This coding decision was based on the low population prevalence of ADHD, OCD, and related conditions (estimated at 1–4% in the general population ^8–10^), such that treating unknown control status as absent introduces minimal bias while preserving the largest possible sample size. Sites that no longer contained both cases and controls following these exclusions were also removed.

To evaluate the impact of each confounder, we compared the TS effect estimate before and after covariate adjustment, including the absolute and percentage change in the beta estimate. Brain regions that survived FDR correction in the unadjusted model were retested in adjusted models using FDR correction restricted to these regions. Regions that remained significant were considered robust to the respective confounder.

#### Genetic pleiotropy analysis

To further investigate potential mechanisms linking brain structure and genetics, we used summary statistics from the latest TS genome-wide association study (GWAS), which included 13,247 cases and 536,217 controls ^11^, along with data from the ENIGMA consortium’s GWAS on cortical surface area, thickness, and subcortical volumes ^12,13^. This analysis explored genetic pleiotropy, where the same single nucleotide polymorphism (SNP) affects both TS/CTD and brain structure, using the SNP Effect Concordance Analysis (SECA) tool ^14^. First, we aligned SNP effects across GWAS summary results to the same effect allele and used PLINK for linkage disequilibrium (LD) clumping to extract a subset of independent SNPs, following parameters from the original paper. Pleiotropy analyses were conducted using an exact binomial test. By focusing on SNPs associated with P1 ≤ {0.01, 0.05, 0.1, 0.2, 0.3, 0.4, 0.5, 0.6, 0.7, 0.8, 0.9, 1.0} in dataset1 (risk factors), we used the R statistical package to perform binomial tests for an excess of SNPs associated in both datasets when P2 ≤ {0.01, 0.05, 0.1, 0.2, 0.3, 0.4, 0.5, 0.6, 0.7, 0.8, 0.9, 1.0} in dataset2 (brain alteration rates). For each of the 144 SNP subsets, binomial tests for global pleiotropy were performed. Permutation testing created uncorrelated datasets by shuffling observed SNP effects and P-values, then repeating the analysis of the 144 SNP subsets. Permuted P-values were estimated for the observed number of binomial tests exhibiting global pleiotropy between the two traits.

### Supplementary Results

#### Identifying Brain Structure Differences in TS/CTD versus Controls through Meta-analysis

In the full-sample meta-analysis, we identified significant associations with TS/CTD in 7 surface area regions, 13 cortical thickness regions, and 2 subcortical volume regions (Supplementary Table S7). In the pediatric group, significant associations were observed in 7 surface area regions, 11 cortical thickness regions, and 2 subcortical volume regions (Supplementary Table S8). All regional findings identified in the full and pediatric samples were replicated in the mega-analysis. In contrast, no significant regional differences were detected in the adult group (Supplementary Table S9). Correlation analyses demonstrated strong agreement with the primary mega-analysis across all groups, including the full sample (r = 0.89, P = 3.56×10^-54^), pediatric sample (r = 0.90, P = 1.62×10^-57^), and adult sample (r = 0.93, P = 2.43×10^-70^).

### Supplementary Figures

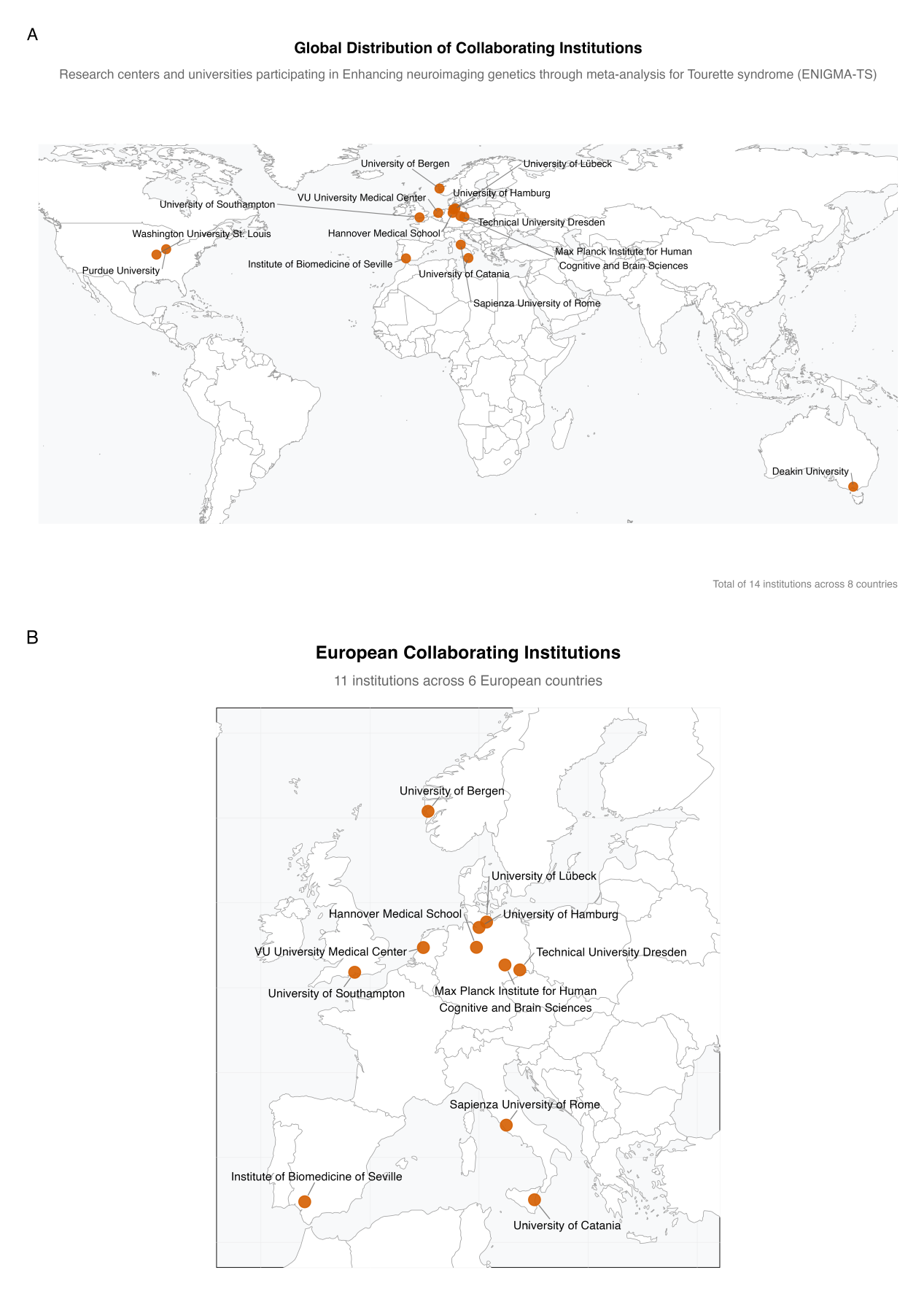

#### Figure S1: Global Distribution of Research centers and universities participating in Enhancing neuroimaging genetics through meta-analysis for Tourette syndrome (ENIGMA-TS)

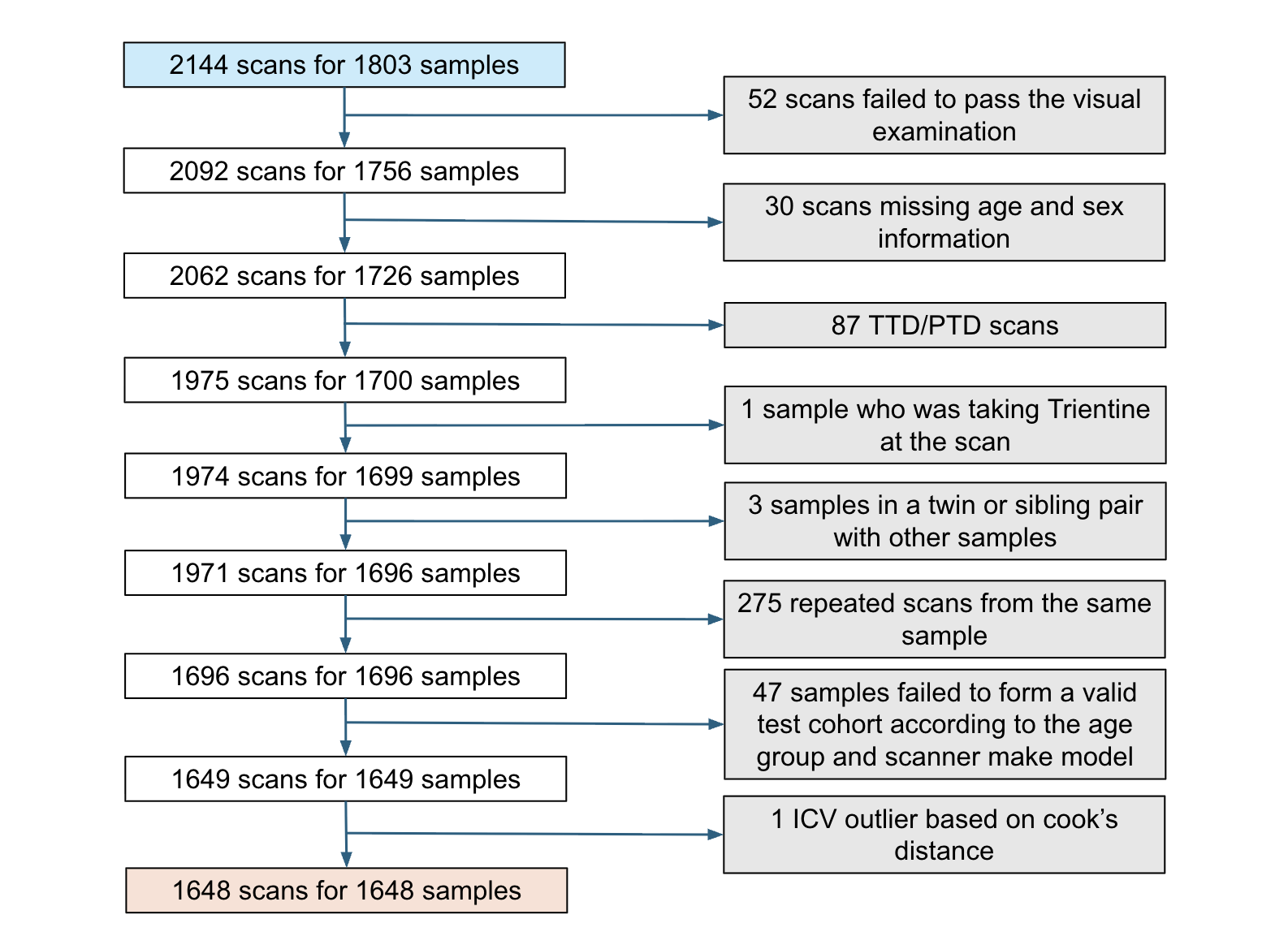

#### Figure S2: Quality Control Workflow – A flowchart illustrating the selection process and subset of participants included in the analysis.

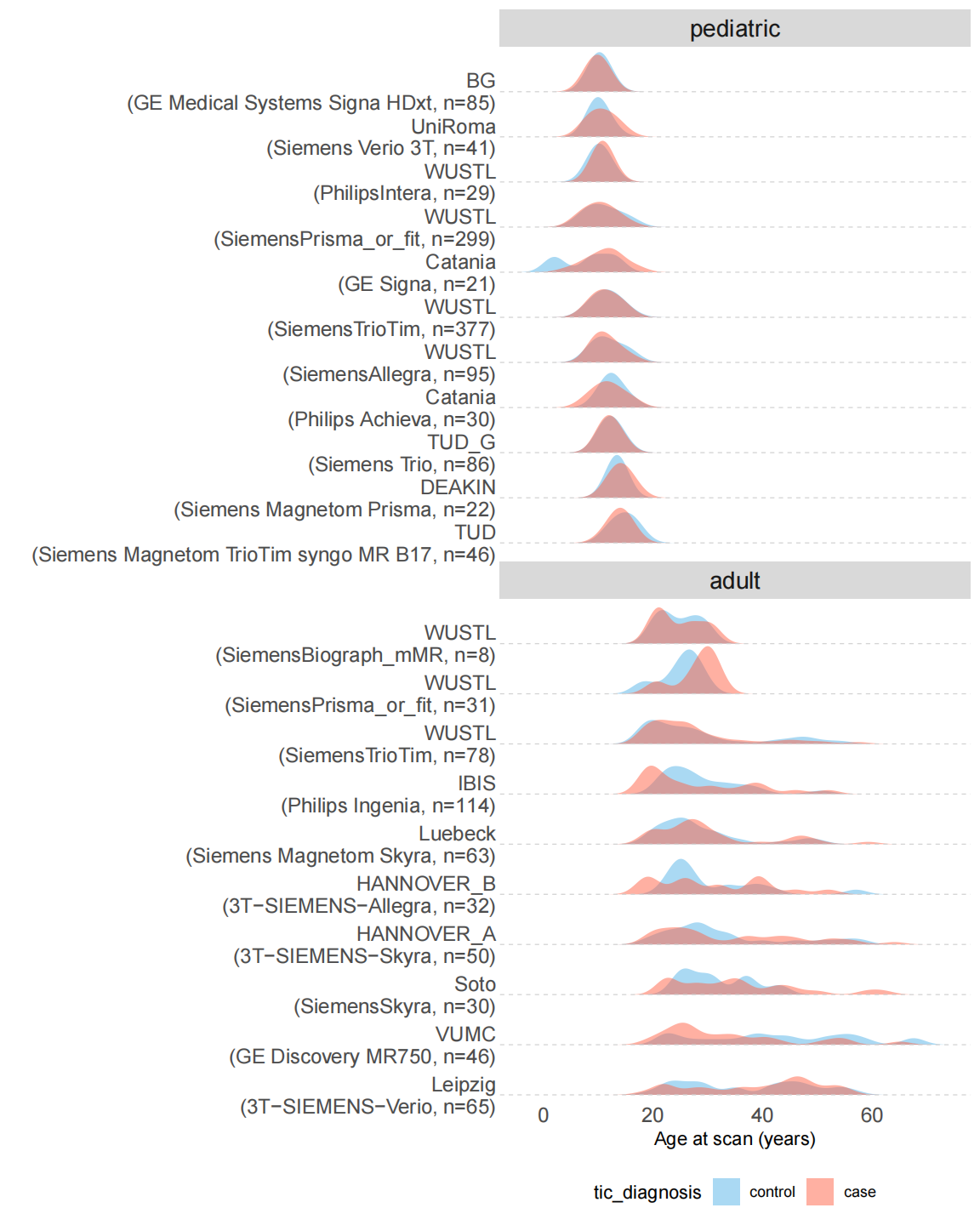

#### Figure S3: Age distribution of TS/CTD cases and controls across all datasets.

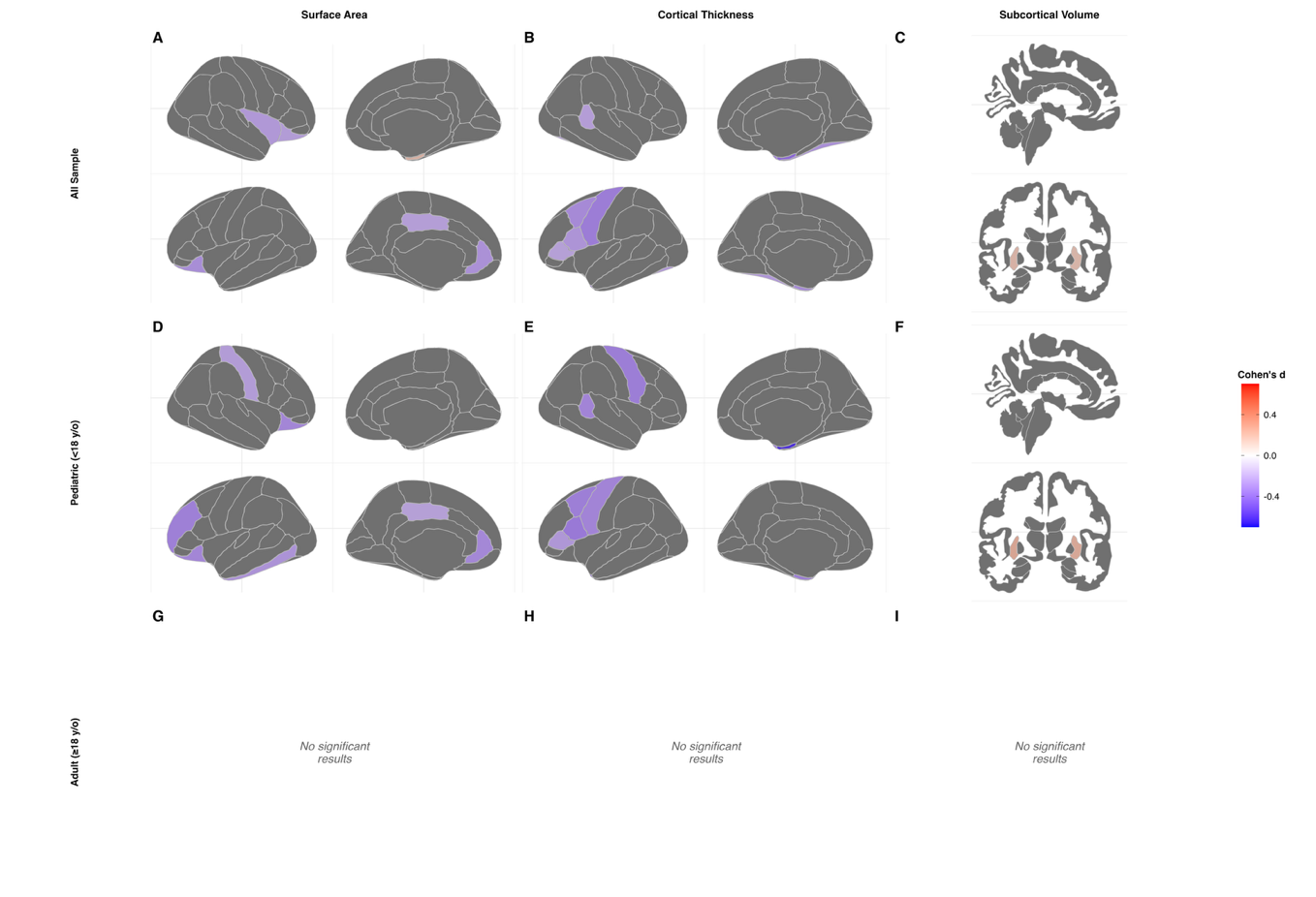

#### Figure S4: Brain maps illustrating effect sizes (Cohen's d) of structural differences between individuals with TS/CTD and healthy controls through meta-analysis.

Only regions reaching FDR-corrected significance (p < 0.05) are displayed. Non-significant regions are shown in dark grey. Color intensity reflects the magnitude and direction of the effect: red indicates larger brain volume/thickness/area in TS/CTD relative to controls (positive Cohen's d), and blue indicates smaller measurements in TS/CTD relative to controls (negative Cohen's d). Each panel shows cortical maps for the right hemisphere (top row) and left hemisphere (bottom row), with lateral (outer) and medial (inner) views displayed for each hemisphere.

(A–C) All ages combined: surface area (A), cortical thickness (B), and subcortical volume (C). (D–F) Pediatric participants (<18 years): surface area (D), cortical thickness (E), and subcortical volume (F).

(G–I) Adult participants (≥18 years): surface area (G), cortical thickness (H), and subcortical volume (I).

*No significant results* indicates that no brain regions survived FDR correction in that group and measure.

A

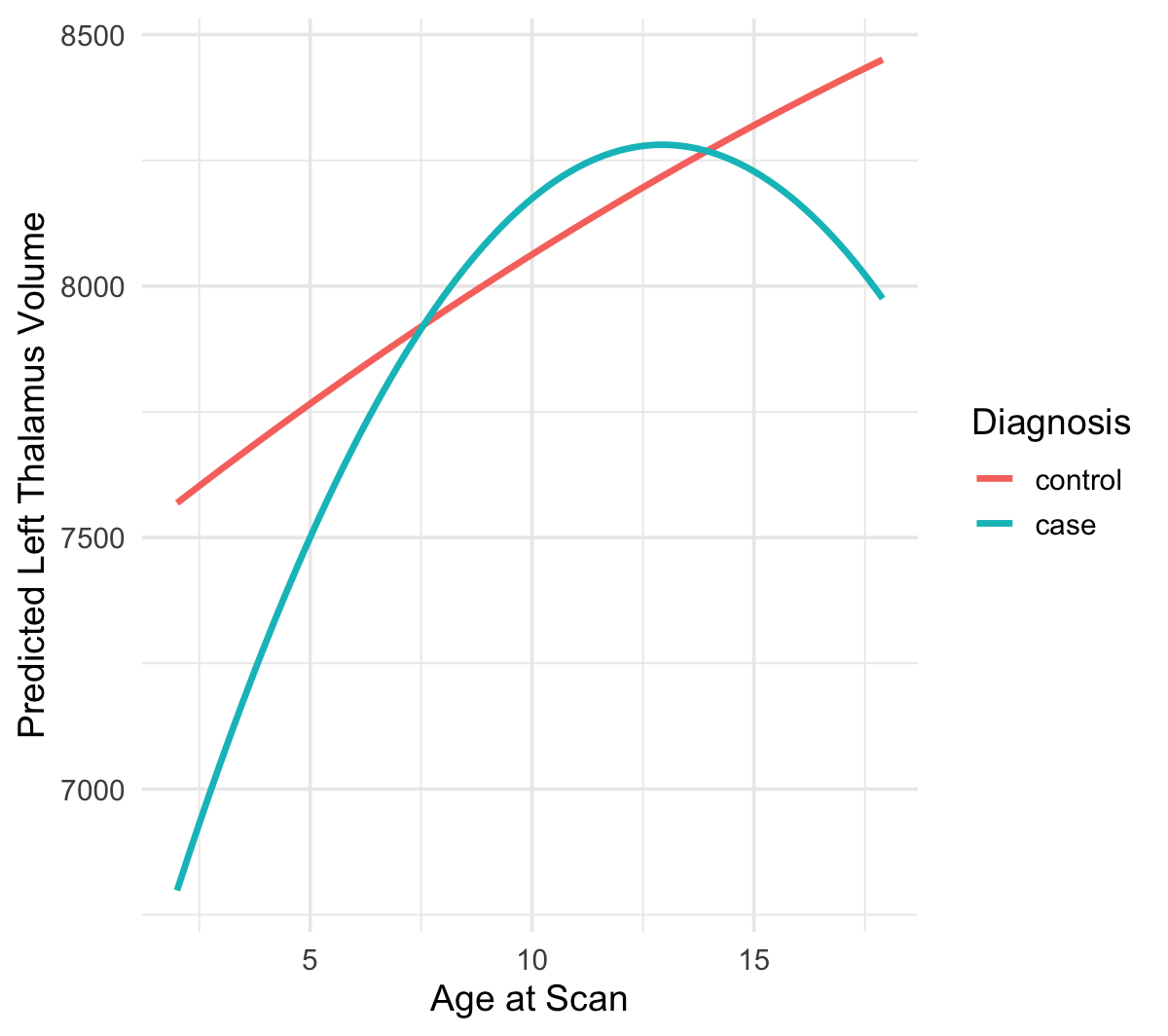

B

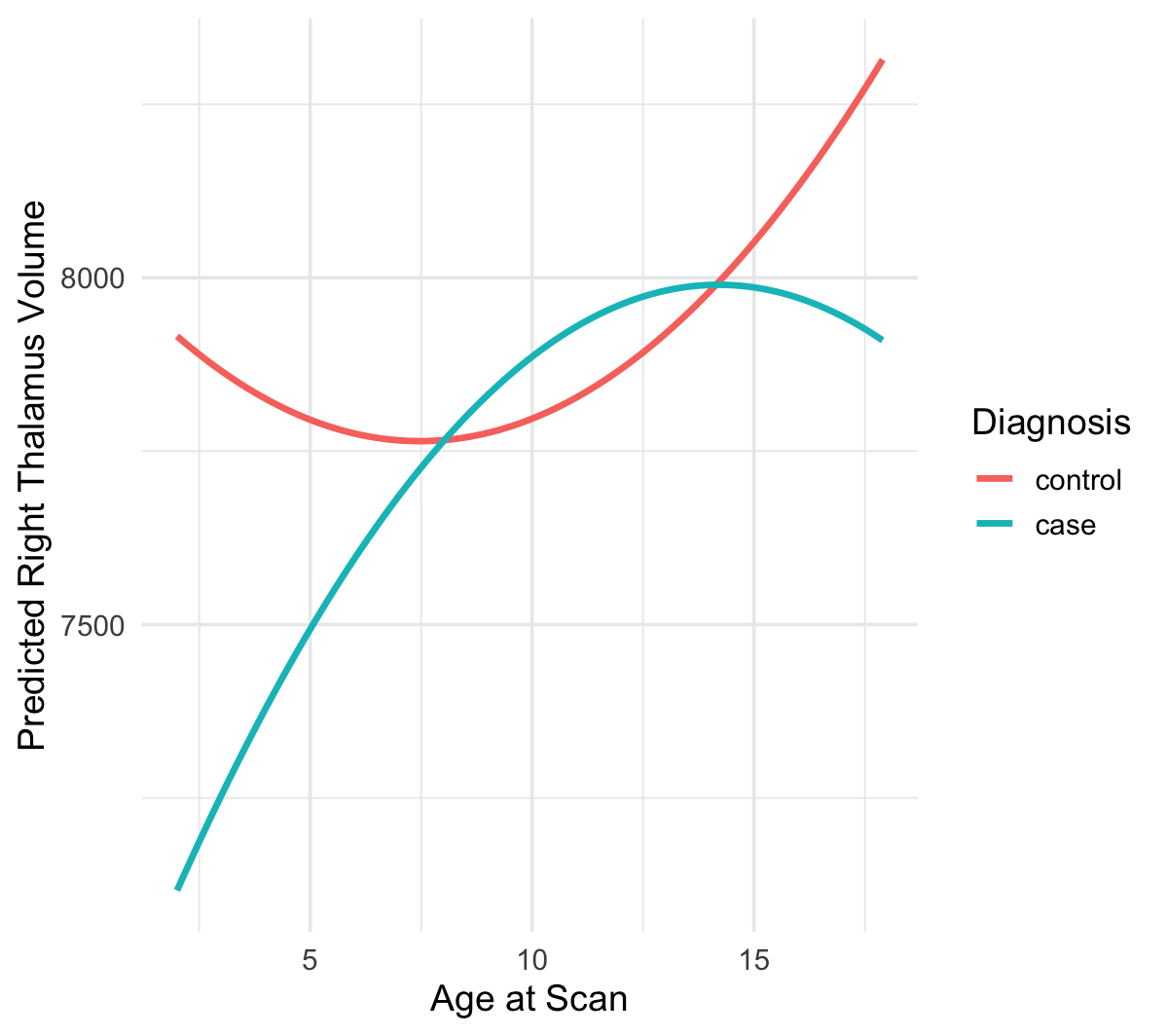

#### Figure S5: Model-Based Predicted Thalamus Volume Across Age by Diagnosis.

The lines represent model-based predictions from a linear mixed-effects model including diagnosis-by-age (linear and quadratic) interaction terms, with sex and intracranial volume (ICV) included as covariates. We excluded sex-by-age interaction terms from the final model to simplify interpretation and plotting, as these interactions were not statistically significant in our analysis. The prediction grid was built using a sequence of ages, crossed with each diagnosis group and adjusted by sex, while holding ICV constant at the group mean. The smooth curves represent predicted brain volume as a function of age, allowing for visualization of the diagnosis-by-age interaction effect while adjusting for covariates. A) left thalamus B) right thalamus.
